# Enhancing local public health decision-making: Incorporating end-user perspectives into influenza forecasting models

**DOI:** 10.1101/2025.08.19.25334039

**Authors:** Rochelle L. Frounfelker, Kareem Hargrove, Katherine Blomkvist, David Rea, Thomas C. McAndrew

## Abstract

**Background:** Influenza has a significant impact on morbidity and mortality, with disproportionate impact on non-White populations. Forecasts of flu timing/intensity have the potential to reduce morbidity, mortality, and health disparities by supporting decision-making by public health officials and clinicians. However, uptake and use of forecasts on local levels is low, with limited communication between model developers and end-users. The goals of this study are to: 1) understand the seasonal flu intervention decision-making process from the perspective of local public health officials and health care providers; and 2) identify these stakeholders’ data needs and priorities for flu forecasting models.

**Methods:** This mixed methods study included a brief survey and two rounds of focus groups with local public health officials and clinicians in a mid-sized metropolitan area in the Northeast US (N=16). Authors used descriptive statistics to analyze survey responses and content analysis to analyze qualitative data.

**Results:** Participants described a decision-making process that included using data from forecast models and other sources to inform health interventions, health communication, and resource allocation. Primary outcomes for decision-making included disease prevention and health care preparedness. Participants articulated a variety of ways that forecasting models could assist them in delivering evidence-informed public health and clinical services, and data needs including sociodemographic characteristics and surveillance information at smaller spatial scales. There was a desire for functionality of models that reduced the time spent gathering information.

**Conclusions:** Findings support the need for a participatory modeling approach to the design of influenza forecasts that improves uptake by including the goals and desires of end-users.

## Background

In the US, influenza (flu) causes 9m-41m infections, 140k-710k hospitalizations, and 12k-52k deaths annually [1]. Of the estimated $11B in total flu-related healthcare costs, $3.2B has been attributed to direct medical costs [2,3]. Flu disproportionately impacts those who are Black and Hispanic [1,4]. Compared to those who identify as White, Black and Hispanic children have a higher probability of worse outcomes due to flu, and Black individuals under 65 have a higher probability of hospitalization and ICU care due to flu [5]. Despite an increased effort to vaccinate against influenza, significant healthcare costs remain.

The Centers for Disease Control and Prevention (CDC) launched the Epidemic Prediction Initiative (EPI) with the goal to improve the science of epidemic forecasts by facilitating forecast projects [6,7]. Projects like FluSight, part of EPI, compile forecasts of incident hospitalizations due to flu to disseminate to public health officials (PHOs) at national, federal, and state levels [6,8,9]. Forecasts of flu timing/intensity have the potential to reduce morbidity, mortality and health disparities by supporting public health emergency response/preparedness [10–12].

Generating forecasts with high performance does not guarantee that they are used by end-users (i.e., public health officials and clinicians) who make policy and programmatic decisions related to flu interventions. Uptake of forecasts at the federal level is high; however, uptake at smaller, more local levels is low because forecasts do not often reflect context-specific constraints/priorities [13]. Although there are exceptions [14], epidemiological modeling can be characterized as being very “siloed,” with limited communication between model developers and end users [15]. There is minimal information on how end-users perceive and actually use models in decision-making [15]. The few studies that assess the relationship between end-users and infectious disease forecasting models highlight lack of familiarity, trust/comprehension of models, and communication between end-users and modelers [13,16].

Forecasting models are too often built without the involvement of public health officials and clinicians, and there is a need to develop connections between computational epidemiology modelers and model end-users [13, 15, 17–19]. This is critical, as communication and inclusion of end-users in the modeling process may better support evidence-informed public health interventions. Public health officials have explicitly asked for better tools to integrate forecasting into flu decision making [6, 20]; however, they are rarely provided the opportunity to prescribe the components of these tools [21].

An initial step to facilitate partnerships between modelers and end-users and develop forecasting tools that address end-users priorities is to undertake a needs assessment to explore what data is currently used and desired by public health officials and health care providers to make evidence-informed policy, programmatic, and clinical decisions. More broadly exploring the decision-making process of end-users and their responses to the seasonal flu can identify information gaps as well as features and functionality of forecasting models that could address these gaps. This mixed methods study takes this approach and focuses on public health and clinical stakeholders in a mid-sized metropolitan area in the Northeast US. The goal of this study is to: 1) understand the seasonal flu intervention decision-making process from the perspective of local public health officials and health care providers; and 2) identify these stakeholders’ data needs and priorities for flu forecasting models.

## Methods

### Ethics

This study was reviewed in accordance with the Declaration of Helsinki and approved by the institutional review board of X university (blinded for review).

### Setting and participants

Study participants were health care providers and public health officials working in a metropolitan area in Pennsylvania interviewed in 2023 and 2024. Eligibility criteria included being aged 18 and over and knowledgeable about influenza prevention and intervention specific to this metropolitan area. A total of 16 individuals participated in two rounds of focus groups (for a total of 8 meetings) (see Table 1). The focus groups were stratified by stakeholder groups: infectious disease physicians, non-infectious disease clinicians, and public health officials working with one of two local health bureaus (4 expert groups total).

**Table 1.** Participant Sociodemographic Characteristics (n=16)

|  | N (%) |
| --- | --- |
| **Gender** |  |
| *Female* | 12 (75.0) |
| *Male* | 4 (25.0) |
| **Employer** |  |
| *Allentown Health Bureau* | 5 (31.3) |
| *LVHN* | 7 (43.8) |
| *Bethlehem Health Bureau* | 4 (25.0) |
| **Highest education** |  |
| *High school* | 1 (6.0) |
| *College* | 4 (25.0) |
| *Masters* | 4 (25.0) |
| *Doctorate* | 7 (43.8) |
| **Clinical role (if applicable)** |  |
| *Pharmacist* | 1 (8.3) |
| *Physician* | 6 (50.0) |
| *Nurse/nurse practitioner* | 4 (33.3) |
| *Registered dietitian* | 1 (8.3) |
|  | **Mean**<br>**(SD)** |
| **Age** | 51.8 (9.5) |
| **Years of experience in**<br>**infectious disease** | 15.8 (13.5) |

### Procedures

Study recruitment began October 1, 2023 and concluded March 31, 2024. Researchers had pre-established relationships with leaders in infectious disease prevention and intervention who work in health care and public health bureau settings in the community. Researchers invited these individuals to participate in the study, and used snowball sampling to reach out to potential participants that were colleagues and met study criteria. Researchers obtained verbal informed consent from individuals prior to enrollment. Participants were offered a $30 gift card to compensate them for their time. All participants declined compensation.

Researchers used a semi-structured moderator guide for focus groups (see Supplemental File 1). Two rounds of focus groups were conducted with each group that worked in the same health care or health bureau setting (4 settings total). Focus groups were co-facilitated by the lead author and senior author (an influenza model developer) and carried out in a private location convenient for study participants. Focus groups lasted between 45 and 70 minutes. The first round of focus groups explored seasonal flu interventions, outcomes of interest to decision making, information currently used to make decisions about flu interventions, and data needs. Prior to the second-round focus group, participants completed a brief survey assessing their preferences and priorities for flu forecasting models. The second round of focus groups built on the first-round focus groups by exploring stakeholder views and use of forecasts, and tools for forecasting; and by asking participants to rank (from 1=most important to X=least important) the spatial levels and populations for which forecasts of seasonal influenza could be most important. Participants were allowed to add items they felt were missing and include them in their ranking. Focus groups were audio-recorded and transcribed by members of the research team prior to analysis.

### Data analysis

Researchers used descriptive statistics to characterize participant responses on quantitative survey questions. Survey questions that asked participants to rank the importance of different kinds of flu forecasts were summarized by calculating the mean rank score for all participants, stratified by end-user group (public health official or health care provider).

Researchers analyzed qualitative data using content analysis with an inductive open coding approach to identify topics emerging from the data [22, 23]. The process began with three coders coding two focus groups independently prior to meeting to discuss results. The analysis team met to discuss codes and discrepancies until consensus was reached. An initial coding structure, including categories and codes, was developed and a codebook, complete with definitions and examples of inclusion/exclusion criteria was created. For instance, the category “Data wants” included subcodes of “vaccinations,” “sociodemographics,” and “zip code.” Analysts used this codebook as a guide for coding the remaining focus groups. The codebook underwent additional refinement as new codes were added, restructured, or changed based on team discussions. After triple coding the first two focus groups, analysts split up and coded the remaining transcripts, coming together to discuss and resolve discrepancies. Quotes which best captured these findings were included in this manuscript. Data were managed and analyzed in MAXQDA mixed methods software [24].

## Results

The model of seasonal flu decision-making for local infectious disease clinicians and public health officials can be characterized as containing four components: information, decisions, actions, and outcomes (see Figure 1).

**Figure 1.**
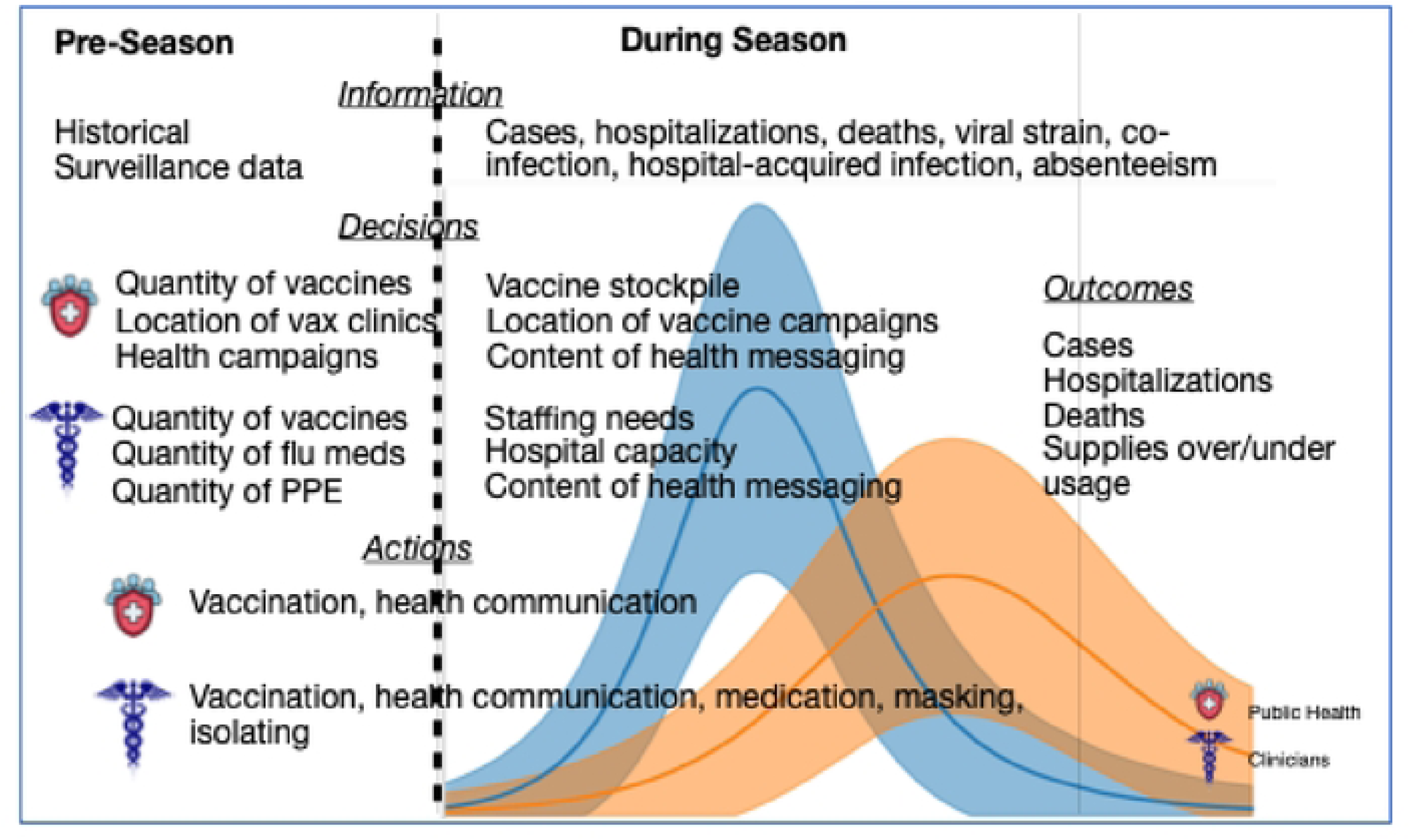
Seasonal Flu Decision-making.

Participants articulated two primary outcomes they worked towards during a typical flu season – disease prevention and health care preparedness. Prevention outcomes that were listed by participants include primary prevention (fewer cases of flu) as well as secondary and tertiary prevention (fewer hospitalizations, fewer deaths, less severe symptoms of flu) in the general population. The importance of these two primary outcomes were emphasized when participants were asked what a “successful” flu season would look like. One public health official summarized, “more people getting vaccinated and less people being sick, fewer people being hospitalized.” Specific to health care preparedness, participants articulated that they need adequate, but not wasteful, amounts of vaccines, medication, and supplies (PPE, hospital beds) to respond to influenza burden during the season, as well as, within the hospital system, staffing to respond to an influx of patients with flu-related symptoms. One infectious disease clinician defined success for both patients and practitioners as “not overwhelming our ICU, having enough PPE for our staff.”

Participants use a wide range of information to make decisions in preparation for, and during, a flu season, to achieve the above successful outcomes. Most participants rely on historical trends and current data on number of cases, hospitalizations, and deaths obtained from state and national-level surveillance systems and data from Pennsylvania Department of Health and Centers for Disease Control. Of the two, one participant noted that data from the Pennsylvania Department of Health was more useful because it provided information on smaller spatial scales and closer to real-time than the CDC. Clinicians also use point of care and health care network data to manually investigate hospital acquired infections, outbreaks in long-term care facilities, co-infection, and health care personnel absenteeism related to the flu. A health care provider explained, “doctor’s offices, ambulatory care places…capture [influenza] because so many of these practices are…within our network…Right now most people are getting flu, RSV, and COVID testing. When…you attend a hospital, or when you’re supposed to, those metrics are very easy for us to access.”

Participants identified priorities related to the information they wanted to inform their actions. For weekly seasonal flu targets, clinicians ranked incident hospitalizations as most important, and incident deaths as least important. In contrast, out of the four options, public health officials ranked incident hospitalizations and vaccinations as equally, and most, important. Incident cases and incident deaths were tied and ranked least important. In terms of forecasts, clinicians ranked incident cases of flu over the typical flu season as most important, and subtype specific incident cases of influenza least important. Public health officials prioritized information on the peak time and peak intensity of influenza during the flu season as more important than incidence cases; similar to clinicians, forecasts on specific subtypes were least important. For both public health officials and clinicians, information on the trajectory of total incident cases was preferable to cases stratified by influenza type. Finally, participants felt that forecasts of cases, hospitalizations, and deaths were more valuable to have when planning vaccine campaigns compared to previous estimates of vaccine coverage (see Table 2).

**Table 2.** Forecasting model priorities of local public health officials (N=9) and clinicians (N=6)

| Question | Clinician Mean Rank Score | Public Health Mean Rank Score |
| --- | --- | --- |
| *Rank the importance of these weekly seasonal influenza targets from 1 (most important) to 4 (least important)* |  |  |
| Incident hospitalizations | 1.8 | 2.4 |
| Incident cases | 2.2 | 2.6 |
| Incident vaccinations | 2.7 | 2.4 |
| Incident deaths | 3.3 | 2.6 |
| *Rank the importance of these forecasts when making flu intervention decisions from 1 (most important) to 6 (least important)* |  |  |
| Incident cases of influenza over the typical flu season | 2.0 | 3.1 |
| Peak time and peak intensity of influenza during flu season | 2.2 | 2.8 |
| Location (district, county, zip code, etc.) of highest intensity for the season | 3.0 | 3.3 |
| Location (district, county, zip code, etc.) of highest cumulative number of cases for the season | 3.8 | 3.3 |
| Location (district, county, zip code, etc.) of highest vaccine coverage | 4.2 | 3.9 |
| Subtype specific incident cases of influenza (e.g., H1 vs. H3, Victoria vs. Yamagata, etc.) | 5.5 | 4.6 |
|  | **N (%)** | **N (%)** |
| *Which of the following two kinds of forecast information are most important for you to have when planning targeted vaccine campaigns?* |  |  |
| Trajectory of incidence cases in total (influenza A and influenza B combined) | 6 (100) | 6 (67) |
| Trajectory of incidence cases stratified by type (influenza A separate from influenza B) | 0 | 3 (33) |
| *Which of the following two kinds of forecast information are most important for you to have when planning targeted vaccine campaigns?* |  |  |
| Forecasts of cases, hospitalizations, deaths presented at a weekly cadence during flu season (MMWR40-20) | 5 (83) | 6 (67) |
| Previous estimates of vaccine coverage | 1 (17) | 3 (33) |
| *Is it important to estimate the reproduction number for the flu season?* |  |  |
| Yes | 6 (100) | 6 (67) |
| No | 0 | 2 (22) |
| Missing | 0 | 1 (11) |

Public health officials and clinicians used the information described above to guide actions related to health care interventions, health communication with the public and patients, and appropriate resource allocation. When asked how forecasting data could benefit their work, public health officials spoke about their mandate to provide vaccines to vulnerable populations and using health communication campaigns to promote, throughout the flu season, prevention initiatives like vaccination. One individual explained,

> “Part of our stance of trying to be that reliable data source for the city of X and Y County as far as being that person to say, like, “why should you get the flu vaccine?” Well, if the message is coming from a public health department, “this is why you should get the flu vaccine” and [we] have good, valid, reliable data …our incentive is to…keep as many people healthy as possible and get good information out to all of the stakeholders in our area. So it’s important for us to have those numbers.”

In contrast to public health officials, clinicians focused on providing medication (in addition to vaccines) and recommending masking and isolation for patients who were vulnerable or sick. One clinician stated,

> “Week to week (surveillance data) helps us to see what’s going on in the community, the number of cases, positivity rate, and it helps you to make medical decisions. Like if somebody in the family is positive and the next member comes in and is seen. I don’t need to be testing everybody so I can make decisions [about testing] based on my number of cases.”

### Forecasts can support public health and clinical efforts

Both public health officials and clinicians discussed ways in which flu forecasts could be used to inform health communication with the public (including patients). Although health bureaus did not have the power to mandate vaccinations or interventions, like isolation and masking, they saw the value that data (either real-time or forecasts) can add towards developing trust. One person commented, “The more information we have, we can leverage to strengthen partnerships…It’ll open doors for discussion and they can do with that what they want to do with it. I don’t know how they’d [businesses, schools] react from an operational standpoint, but having that piece of information helps us just be better partners to them.” In contrast to public health officials who focused on corporations and businesses, clinicians talked about building trust with individual patients and families. One clinician noted, “Surveillance data helps us the next season with the story we tell our patients. Last year we had a lot more hospitalizations and pediatric deaths. So you really should get your kid a flu shot this year. That kind of stuff can be helpful.”

When discussing how forecasts can improve care, clinicians focused on improving resource allocation while public health officials focused on identifying at risk populations in their jurisdiction. Clinicians talked about using forecast data to plan for resource allocation in the health care system and determine if they needed to order more vaccines or medication to prepare for a more severe flu season. An infectious disease clinician stated,

> “I can think of a couple of areas where we would benefit [from forecasts]. So, if we see a bad season coming potentially staffing in the hospital. Vaccines—so, we tell our patients to get vaccinated around October, predicting that it will come sometime in December. And we want everyone to be vaccinated at least two weeks before the season begins. So do we adjust that? So, that could be not just the hospital systems but pharmacies for anyone who is providing the flu vaccine. The flu drives that we do might change the date a little bit, based on that. Our availability to have certain medications, you know, sticking for the pharmacies, like Tamiflu might be adjusted based on the timing of when the flu season starts and how bad it’s going to be.”

With a greater focus on primary prevention, health bureaus discussed using forecasting data to identify at risk populations and plan accordingly. A public health official explained,

> “I think the vaccinations, the location of where people are getting their vaccinations, I think is really key to our strategic plan here, because if we’re finding that most people are getting their vaccines [somewhere] other than here [the local health bureau], then we have to reevaluate whether or not that’s a function that we want to continue for the general public, or if we want to… move… our medical van… into the pockets of the underserved.”

### Barriers to influenza decision-making

Participants highlighted gaps in information to make evidence-informed decisions that they hoped forecasting models could address. Infectious disease clinicians articulated wanting past, current, and forecast data in one place. One person stated, “In an ideal [platform] we would see the forecast and we would also be able to see prior data too. A second participant added that forecast data in addition to real-time data can communicate trust in modeling. They said “I think if you show forecasted data compared to what actually happened that in itself is important information too. And if it’s reliable then people will trust it.” Finally, clinicians want both counts and rates of infections to compare severity to past seasons. An individual explained “just the number is not enough. Like, I need to know that it’s the rate, and compared to prior years, is different. So, that representation of the data would alert me that there’s something wrong.” Participants highlighted the desire for data on smaller spatial scales than was typically available from current forecasting model sites. A health bureau employee explained,

> “Sometimes we use the term OJ – out of jurisdiction. If it’s outside the city limits, we can do very little with it. If it’s outside the county, we can do even less with it. So if there was some reason it was going to peak here two months before, two months after it was going to peak nationally or in California, then I’d rather know what’s happening here.”

Clinicians echoed this desire for more local data, with one person stating,

> “Influenza is extremely regional. I mean you can have significant outbreaks, widespread outbreaks, in one part of the state and not in another, a couple counties over and so on…now we’re such a large network, it’s not a mom and pop shop so we’re responding to county level cases.”

### Preferences and priorities for data and forecasts

Participants identified priorities for the presentation of forecasts, with an emphasis on gathering past, present, and future (e.g. forecasts) information at several spatial scales about cases, hospitalizations, and deaths in a single place. An infectious disease clinician stated, “I guess one thing, and hopefully you guys fix this, is the information in multiple spots, so I could spend 30 minutes to an hour to look at data at the national level, state level, and local level. So I’m…spending a lot of time trying to look at all of the information I need.” Specific to different spatial levels, stakeholders were interested in layering case, hospitalization, and death data on multiple scales and having access to data outside of the forecast platform, such as downloading as a spreadsheet. One person noted, “A filter would be helpful where you could go down to one individual neighborhood, county. But sometimes it is nice to see a few close to each other; I don’t know if that’s asking for too much, but, the ability to check [incidence rate between] three and only these three. I [would] like the ability to download all the data into excel.” Finally, stakeholders were interested in functionality related to presenting visuals for communicating benefits and risks to the population and individual patients. A clinician explained, “A visual of some sort…could be impactful to families. So let’s say the vaccine efficacy is only 40% one year. It’s usually not real high, but if we had a model that…showed families… how much your risk is decreased [if you get vaccinated]. This is what could happen to you if you…get [the] flu because you didn’t get vaccinated.”

When each of the four groups (infectious disease clinicians, other health care providers, and two local health bureaus) ranked data priorities for parameters related to spatial scale, geographical settings, physical health issues and conditions, and sociodemographic characteristics, there was some consistency. The most important information for which all groups were unanimous was: stratifying by age, pregnancy status, and past respiratory illnesses/chronic respiratory disease. Importantly, opinions on the most important spatial level and geographic setting were mixed. For instance, when asked which sociodemographic variable is most important, all four groups ranked age as most important. Interestingly, both groups of clinicians requested to include information on an individual’s social vulnerability index, a metric included in their electronic health record system. All groups identified pregnancy and respiratory illnesses/chronic respiratory disease as the most important physical health issue. There were variations in priorities for spatial scale and geographical settings. Groups were split on prioritizing data from hospitals (both in and outpatient) and area schools. There was no agreement among the groups on the spatial scale that was most important for making decisions, with health bureaus wanting smaller spatial scales (city and neighborhood) than clinicians larger spatial scales (county and state) (see Table 3).

**Table 3.** Group Ranking Data Priorities and Needs (N=4 groups)

| Parameter | Rank | Group Rank |  |  |  |
| --- | --- | --- | --- | --- | --- |
|  |  | *Infectious Disease Clinicians* | *Other Clinicians* | *Health Bureau 1* | *Health Bureau 2* |
| *Spatial* | 1 | County | State | City | Neighborhood |
|  | 2 | City | County | Neighborhood | Ward |
|  | 3 | Zipcode | Zipcode | Zipcode | City |
|  | 4 | State | City | Ward | County |
|  | 5 | District | Ward** | County | Zipcode |
|  | 6 | Neighborhood | Neighborhood** | State | District |
|  | 7 | Ward |  | District | State |
| *Sociodemographics* | 1 | Age | Age | Age | Age |
|  | 2 | Social Vulnerability Index* | Social Vulnerability Index* | Ethnicity* | Race/Ethnicity |
|  | 3 | Race/Ethnicity | Race/Ethnicity | Race/Ethnicity | Sex |
|  | 4 | Sex | Sex | LGBTQ+* |  |
|  | 5 |  |  | Sex |  |
| *Settings* | 1 | Hospitals | K-12 | Hospitals | K-12 |
|  | 2 | Long term care facilities | Factories | Long term care facilities | Long term care facilities |
|  | 3 | Redi-clinics** | Colleges | K-12 | Hospitals |
|  | 4 | K-12** | Businesses | Factories | Redi-Clinics |
|  | 5 | Factories | LTCF | Businesses | Factories |
|  | 6 |  | Hospitals | Redi-clinics | Businesses |
|  | 7 |  | Colleges* |  |  |
| *Physical health* | 1 | Pregnancy | Pregnancy | Pregnancy | Respiratory Disease |
|  | 2 | Cardiovascular Disease | Immunosuppression* | Chronic Illnesses | Pregnancy |
|  | 3 | Diabetes | Obesity | Respiratory Disease | Diabetes |
|  | 4 | Respiratory Disease | Respiratory Disease | Cardiovascular Disease | Cardiovascular Disease |
|  | 5 | Smoking* | Cardiovascular Disease | Diabetes |  |
|  | 6 | Obesity* | Diabetes |  |  |
|  | 7 | Kidney Disease* |  |  |  |
|  | 8 | Liver Disease* |  |  |  |
|  | 9 | Immunosuppression* |  |  |  |
|  | 10 | Malignancy* |  |  |  |

\*Added by participants
\*\*tied

## Discussion

Local public health officials and health care providers described a decision-making process in which data on the burden and distribution of influenza informs actions related to health interventions, health communication, and resource allocation. Stakeholders were aware of and used forecast models as a source of data in their work, and were enthusiastic about working with modelers to address information gaps that could improve their decision-making. Model recommendations included specific data needs as well as model functionality.

Aligned with other research, there was overall positive feedback among stakeholders about forecast models [16]. Our findings that clinical stakeholders see models as an information source for resource allocation, such as ensuring healthcare staffing needs are met and having adequate supplies, including PPE, support other qualitative research on this topic [16, 21]. Results also reinforce that local stakeholders desire models with more fine grained spatial scales that are contextually relevant; state and national level data are not always appropriate for decision-making [21, 25, 26]. Additionally, our findings support that there is a significant gap in surveillance and modeling efforts stratified by sociodemographic characteristics to identify and take action with at-risk communities and groups [21].

This study adds to the literature in several ways. First, prior studies indicate that stakeholder needs and priorities vary based on the unique aspects of their jurisdictions [21]. Our study shows that even within the same jurisdiction, data needs depend on the stakeholder group (public health officials vs health care providers). Second, it is significant that information gaps and priorities are framed within the context of the mission and reach of various stakeholders. Local public health authorities sought data to inform their work on health communication and vaccination campaigns for the most vulnerable and at-risk groups in their community without routine health care. In contrast, health care providers focused more on interventions and resource allocation within a large health care organization. Taken together, this research highlights the importance of context when integrating forecast models into routine practice.

### Limitations

Limitations to this study include a small sample size focused on one metropolitan area in the Northeast US. Findings may not be generalizable to other communities both in the US and abroad. This shortcoming is offset by having rich data on the decision-making process from one location, which may be relevant to other jurisdictions. Additional research with a larger sample carried out in diverse locations is needed. Another potential limitation is that one of the study leads who co-facilitated the focus groups had prior interactions and working relationships with many study participants; this may have led to bias in how participants answered questions. The first author, however, had no prior interactions with participants and led all focus groups. Additionally, these prior working relationships may have promoted more openness to share ideas with the study team.

### Implications

Despite these limitations, this study has important implications for infectious disease forecast modeling. This research shows how the preparation for and mitigation of seasonal flu necessitates a complex process with various stakeholders (clinicians and public health officials) playing different roles. While the process goals are largely universal (keeping the local population healthy while mitigating waste), each stakeholder group within the process faces a different group of constituents; clinicians are primarily concerned about the patients and providers in their health system while public health officials are concerned with the organizations and the community at large. Naturally, they each have different decisions to make each season. The primary decisions of public health officials is how much vaccine to order and to best distribute them to specific groups while the primary decision for clinicians is individual patient care and hospital staffing levels. Public health decisions are largely preventative, coming in the preparedness stage while clinician decisions are largely reactionary occurring during an outbreak. To inform these different decisions, each group reported desiring different types of data and forecasts but neither had easy access to information they felt they needed.

These findings point to the relevance of using a participatory modeling approach to design models to aid in decision-making. Participatory modeling is an approach for engaging stakeholders in the development and use of models designed to aid decision-making in the face of uncertainty [27,28]. The rationale of participatory modeling is that integrating scientific research and local knowledge leads to both better science and decision-making. When experts participate in constructing models, it improves model quality by integrating their experience and understanding of broader social contexts otherwise not captured by scientists [29]. When experts participate in using models, it facilitates communication between stakeholders, promotes broader support for consensus building and collective action, and enhances end-user capacity to make evidence-based decisions [29]. Our findings indicate that public health officials and health care providers have recommendations and ideas for data needs and functionality of forecast models that can best help them respond to seasonal flu and make evidence-informed decisions. Integrating this input into model development may improve uptake and use of forecasting models by local stakeholders. Canada’s community of practice focused on modeling for public health (mod4PH) is one initiative to promote a participatory modeling approach and has potential for applications related to flu forecasting model development and implementation [30–32].

From the viewpoint of mathematical modeling that aims to support public health and clinical preparedness, there are 3 key takeaways: local-level forecasts may be as, or more, actionable than state-level forecasts, forecasts can be used in a diverse number of ways to positively impact the health and well-being of a population, mathematical models of an outbreak can be better trusted if past and present data are presented along with an ongoing description of model forecast accuracy.

The majority of organized forecasting efforts (e.g. FluSight) for influenza are produced at state-level [8, 9, 33]. These are lengthy endeavors that have changed evidence-based public health decision making. However, data from this study shows that state-level forecasts may not be useful for a local, city-level public health bureau. A focus, for mathematical modeling, on local and city-level public health practice could be fruitful. Traditionally, forecasts are used to report the potential number of incident cases, hospitalizations, or deaths associated with influenza [6, 12, 32, 34, 35]. This work highlights the diverse number of ways that forecasts and modeling can be used to improve the health and well-being of a population. In particular, forecasts could support public health messaging if they are trained to forecast specific age groups, sexes, comorbidities; and forecasts can be used in clinical resource allocation. Public health messaging and resource allocation are novel avenues for the discipline of infectious disease forecasting to pursue. For public health officials and clinicians in this study, trust in models can be increased with transparency about model performance. We recommend that the mathematical modeling community adopt appending to forecasts for the present season, surveillance data from previous seasons (if available) as well as near real-time evaluation of forecast accuracy.

## Conclusion

Flu forecasts can reduce morbidity and mortality related to seasonal influenza by guiding the decisions of public health officials and clinicians. Uptake and use of forecasts on local levels can be improved by developing models that respond to the data needs and priorities of local jurisdictions. Partnerships between modelers and end-users is a promising avenue for making forecasts accessible and useful, and can ultimately promote evidence-based public health decision-making.

## Declarations

### Ethics approval and consent to participate

Ethics approval was obtained from the Institutional Review Board of Lehigh University, reference number 2091581-1. All participants provided written informed consent.

### Availability of data and materials

The datasets used and analyzed during the current study are available from the corresponding author on reasonable request.

### Authors’ contributions

RF and TM designed the study and carried out data collection. RF, KH and KB analyzed the data. RF, DR and TM wrote the manuscript with input from all authors.

## Acknowledgements

We would like to thank members of the X Health Bureau, X Health Bureau, and X Health Care Network (blinded for review) for contributed their time and ideas to this study.

